# Joint contact forces during barefoot, minimal and conventional shod running are highly individual

**DOI:** 10.1101/2024.11.27.24318053

**Authors:** Lena Kloock, Andrea Arensmann, Myriam de Graaf, Meike Gerlach, Kim Boström, Heiko Wagner

## Abstract

The supposed benefits of barefoot running are an often debated topic, with many studies investigating footwear influences on the kinematics and kinetics of running. Few studies, however, have analysed the effect on joint contact forces (JCFs). In this study, we investigated the influence of different footwear on the JCFs of the hip, knee, and ankle during running using a 3D musculoskeletal model. Kinematics were recorded from 16 volunteers while running on a treadmill at two speeds (2.0m*/*s and 2.5m*/*s) either barefoot (BF), wearing minimal shoes (MM), or normal shoes (NS). Alongside the JCFs, stride parameters and joint angles were examined using a generalised linear mixed model. Results showed a decrease in the hip JCF from BF to MM to NS, no consistent changes in the knee and an increase from BF to MM to NS at the ankle. However, these changes mostly had small effect sizes, so it’s unclear how relevant they are. The individual responses were much larger and showed opposite effects, indicating that the effects of footwear are highly individual and probably depend on the running style and characteristics of each runner.

## Introduction

In recent years, there has been a big surge of interest in barefoot and minimalist running. With about 34% (23.7 million) of Germany’s population over 14 years running regularly^1^, the question of whether to run barefoot or shod affects many. People have claimed that barefoot running might decrease injury prevalence by decreasing joint loading^2–5^. There definitely seems to be enough evidence that footwear has an impact on joint kinematics^6–10^and kinetics^9–13^, but the results regarding the joint contact forces (JCFs) are conflicting^3–5,8,14,15^. Reducing the peak joint forces could aid prevention of injuries^16,17^, and disorders such as osteoarthritis^18,19^. Therefore, it is important to better understand how footwear can influence the JCFs during running.

The proposed working mechanism for why barefoot running would negate high joint loads lies in the altered kinematic patterns it induces. When running, substantial impact forces must be absorbed after initial ground contact to reduce the vertical velocity of the individual body segments and prepare for propulsion^20,21^. When wearing cushioned shoes, this impact is reduced, leading to a more comfortable running experience. In barefoot running, this cushioning is absent, and the full impact force is perceived by the runners, who might then adjust their running pattern accordingly in an attempt to minimise the impact forces^22–25^.

Indeed, previous studies have shown that barefoot running influences the kinematics of the lower extremity^7,8,11,26^. It has been shown that barefoot running leads to a shift from a rearfoot strike to a more midfoot (or forefoot) strike pattern, as evidenced by an increased plantarflexion angle at heel strike^2,6,8,12,22,27^and a decreased footstrike angle^7^. This footstrike effect has also been reported in studies comparing running in minimalist versus regular running shoes^7,9,12^. Further along the kinematic chain, most studies found a higher knee flexion^2,3,6,9,28^during shod running. However, not all studies found significant changes in the knee angle^7,8,29^. In the hip, there are even more conflicting results, with some papers reporting unchanged hip kinematics^6,9^, while others reported increased^8^ or decreased^3^ hip flexion angles in shod versus barefoot conditions. However, most studies only investigated a small number of steps, and the study protocols of the various studies differ, with, e.g., some investigating inter-subject effects and others intra-subject effects.

Besides the influence on joint angles, studies have also found an influence of footwear on stride parameters. Most studies indicate that runners show a decreased stride length while running barefoot^6–8,22^, as well as decreased stride durations, contact times and flight times when compared to shod running^30^. These changes in running kinematics will lead to changes in the impact force profile. Increased stride lengths and footstrike angles are associated with an increase in impact load on the body, even when running velocity remained unchanged^15,20,24^. Based on the larger stride lengths that were reported in shod running, one can expect higher impact forces there. Indeed, studies reported larger peak vertical ground reaction forces^22,28^and impact peaks^22,27^in shod vs. barefoot running for habitual rearfoot strikers.

The changes in kinematics and impact profiles also lead to changes in the required muscle activity. Reduced activation patterns for the ankle plantar flexors, knee extensors and hip extensors were found between shod and barefoot running^3,31,32^. This change in muscle activation is related to a redistribution of work from the knee to the ankle in barefoot versus shod running^6^ as well as minimalist shoes versus conventional running shoes^6,12^. These changes in work and muscle activity are expected to have major consequences for the JCFs, as it has been shown that muscle activity is one of the major influences on JCFs^15,33–36^and have a larger effect than changes in shoe midsole cushioning^15^.

In addition to inducing altered running kinematics, Braunstein et al. theorised that shoes might also have a more direct influence on the joint forces,^37^. They posit that thicker shoe soles increase the moment arm between the ground reaction forces and the ankle and knee joint, leading to higher joint torques and muscle forces, and therefore higher JCFs with thicker soles.

Several studies have already investigated the JCFs during running with different shoe types, with the use of musculoskeletal models, but the results are still conflicting. For the hip, some articles report an increase in JCFs from barefoot to shod conditions^3,5^, while others report a decrease^8^. Meardon et al. compared the hip JCF between different types of shoe cushioning and found no difference between harder and softer midsoles^14^, while Thomas et al. found increased hip JCFs with a softer midsole^15^. This lack of consistency is also present in the findings for the ankle and knee JCFs. With shoes and/or more cushioning, some studies reported increasing knee forces^3^, some decreasing forces^14^ and some found no differences at all^8,15^. For the ankle joint, similar conflicting results can be found^8,14,15^. These inconsistencies might again be explained by the differing study protocols, running speeds and shoes.

Additionally, the conflicting results in the literature regarding the JCFs might find their origin in subject-specific responses to footwear interventions^38,39^. Regarding different dependent variables, it has been shown that not every individual reacts in the same manner to different footwear^38,40–42^. Several concepts, like the ‘comfort filter’ or the ‘preferred movement path’^39^, were introduced and might hold an explanation for why individuals react differently. However, so far, all studies regarding JCFs in different footwear neglected the individual response of subjects and only focused on the population means. As this has indeed quite an impact on the interpretation of the results, the individual responses should not be ignored.

Aside from barefoot and conventional shod running, running in minimalist shoes is another trend that has recently gained popularity^23,43^. To our knowledge, there are no papers yet that investigate the JCFs in all three of these trends at different speeds. Additionally, most studies either look at walking speeds or fast running speeds. However, the differences between different shoe conditions are not only relevant for competitive runners and experienced recreational runners but also for casual runners moving at slower speeds or with a less well-developed technique. Since effects might be different for this group, they should not be neglected in running studies.

Therefore, the aim of this study was to analyse the influence of minimalist shoes, normal shoes and barefoot running on the JCFs of the hip, knee and ankle during recreational running velocities. An inverse dynamic model was used which has been validated using *in vivo* measured forces for the knee^44^, as well as for the hip^45^ (see Supplementary Figure S.1). This model allows us to calculate ground reaction forces and JCFs during treadmill running. We hypothesised that, due to the aforementioned changes in the kinematics, namely higher ankle dorsalflexion, higher knee flexion and larger stride length, as well as due to larger moment arms, and higher muscle activity, the peak JCFs will be larger in shod than in barefoot running for all investigated joints. Minimalist shoes fall in between barefoot and normal shoes in terms of moment arm and cushioning, therefore, we expect them to lead to peak joint contact forces that are lower than normal shoes but higher than barefoot across all three joints. As we expect to see inter-individual differences, we additionally analysed the effects of the footwear interventions for each subject separately.

## Methods

### Subjects

A total of 16 subjects (9 female) volunteered to take part in this study. They had an average age of 24.38 *±*4.81 years, an average height of 175.35*±* 6.96 m and an average weight of 71.79 *±*8.79 kg. All subjects were physically active for at least 1.5 hours per week and had no injury to the lower extremity in the last six months that led them to miss their regular sports activities for more than two weeks. Only habitual rearfoot strikers were included in the study. Prior to the start of the measurements, all subjects were informed about the measurement process and gave their written consent. Ethical approval was granted by the local ethics committee of the Department of Psychology and Sport Science, University of Muenster(#2023-36-MvdH-FA), and all experiments were performed in accordance with relevant guidelines and regulations.

### Experimental procedure

Subjects were asked to run on a treadmill (pulsar, h/p/cosmos, Nubdorf, Germany) for three different footwear conditions: barefoot running (BF), minimalist shoes (MM) (Minimus TR, NewBalance, Warrington, UK) and normal running shoes with a large midsole (NS)(Gaviota 4, Hoka One One, London, UK). All three footwear conditions were investigated at two speeds (2.0 m/s and 2.5 m/s), making for a total of six trials. Prior to the start of the measurement, subjects performed a two-minute familiarisation run, where they ran at each velocity for one minute in the first shoe condition. During each trial, subjects were asked to run for five minutes, while wearing an IMU-based motion capture suit (XSens MVN Link, Movella, Enschede, Netherlands). Whole-body kinematics were recorded using the Xsens suit at 240Hz during the last two minutes of the trial, to ensure participants had gotten accommodated to the new footwear. As a result of the motion capture process, both velocities of a footwear condition were performed consecutively, before moving on to the next condition. The order of the footwear conditions, and the order of the velocities within the footwear condition, were randomised. In between trials, the subjects got a minimum break of five minutes, that was prolonged if the subjects were not sufficiently rested, as indicated by a Borg RPE score higher than 9, where 6 indicates total rest and 20 complete fatigue^46^.

### Biomechanical model

To calculate the JCFs, the biomechanical model Myonardo® (version 7.2.0, Predimo GmbH, Münster, Germany) was used^44^. Myonardo was developed in MATLAB using the Simscape Multibody 3D simulation environment (Version 2023a, The MathWorks, Inc., Natick, Massachusetts, United States). This whole body model consists of 23 segments, 23 joints with 66 degrees of freedom, and 692 muscle-tendon units. The mass and inertia of each segment were scaled relative to the body mass and height^47,48^. The kinematic data were filtered with a 6 Hz lowpass filter first order before being imported into the Myonardo, which calculated an estimation of the ground reaction forces and the JCFs at 120 Hz^49^. The model was validated by comparing measured JCFs from instrumented knee endoprosthetics with JCFs calculated from simultaneously recorded kinematics during walking and squatting^44^. In the attachments, you can find additionally a validation of the hip joint, using endoprosthetic data recorded during one-legged stance^45^.

### Data analysis

Data analysis was performed in MATLAB (Version 2023a, The MathWorks, Inc., Natick, Massachusetts, United States). In order to extract values for each stride, the treadmill data was separated into individual strides that were defined from heelstrike to heelstrike of the same foot. All dependent variables were calculated per stride of the ipsilateral stride side (e.g. left hip joint force was calculated from left heel strike to left heel strike). As no effect of body side was suspected, data for both body sides were combined leading to a mean of 311.18 ±31.95 evaluated strides per subject, per condition, and per speed. The footstrike angle was calculated as the angle between the ground and the foot at initial contact. For the JCFs, the peak value from each stride was calculated per joint. For a better comparison, each peak was normalised to each person’s body weight. The flexion angles of the individual joints were taken at the time of the peak JCF of that joint. For the joint angles, the T-Pose is the 0° reference, so a negative hip flexion corresponds to a forward movement of the upper body relative to the thigh (i.e., anteflexion), a positive knee angle indicates knee flexion, and a positive ankle angle corresponds to plantarflexion. To investigate individual changes, the relative increase in the normalised JCFs was calculated for each of the three possible footwear comparisons.

### Statistical analysis

To statistically evaluate the experimental data, we chose to use a generalised linear mixed model (GLMM) analysis rather than a traditional repeated-measures analysis of variance (ANOVA) for several reasons: First, our data are unbalanced, as participants had different numbers of observations (‘strides’ in our case) across subjects and conditions. GLMMs can effectively model unbalanced data by estimating individual-specific variances and covariances, ensuring a more accurate representation of variability within the data set. Second, our data set had a nested structure, with individual strides nested within the participants, represented by the ‘subject’ variable. GLMMs are well suited to handle this type of data by incorporating random effects that capture individual variability within nested groups. In addition to random intercepts, we also included random slopes, which further improved the goodness of fit. Finally, GLMMs are much more robust to non-normally distributed data, since all that matters is that the residuals are normally distributed.

An important prerequisite for a successful and reliable statistical evaluation is the exclusion of outliers, which are notorious for experimental data, especially as in our case they result from model simulations based on measured kinematic data. For this analysis, any point outside 1.5 times the (0.25, 0.75) interquartile range of each condition was considered an outlier and removed before entering the GLMM analysis.

The data were fitted using MATLAB’s `fitglme` function, selecting the maximum pseudo-likelihood fit method, effects coding, normal distribution family with identity link function, and the following formula

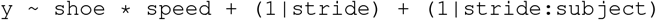

where `y` is the dependent variable. For the analysis of individual subjects, we performed one linear model fit per speed according to the formula

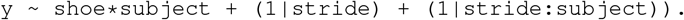

That is, we treated the subject variable as a fixed effect and kept the strides as a random variable nested within the subject variable. In each of these cases, we analysed the data with respect to eight different dependent variables: JCFs and flexion angles for the hip, knee and ankle, plus the footstrike angle and stride length. For each dependent variable, we performed a separate GLMM fit and subsequent calculation of fixed and random effects as well as post-hoc pairwise comparisons between footwear conditions. For the former, we used MATLAB’s `anova` function (which performs a type-III ANOVA on the outcome of the GLMM fit) and, for the latter, we used the `emmeans` and `contrast_wald` functions from the ‘emmeans’ package. Effect sizes were calculated as partial eta squared (*ph*^2^) and interpreted according to Cohen’s rule of thumb with the limits [0.01, 0.06, 0.14] for small, medium and large effect size, respectively^50^.

To validate our GLMM analysis, we generated diagnostic plots to assess whether the distribution of residuals was consistent across predictor variables (homoscedasticity) and to ensure that residuals followed a Gaussian distribution, using Q-Q plots and histograms.

## Results

### Stride parameters

The stride parameters footstrike angle and stride length increased when comparing barefoot running with shod running in both speeds.

With a closer look at the differences within a speed, footstrike angle showed significant increase from BF to MM to NS in both speeds (2.0: BF to MM: *p* < 0.001, *ph*^2^ = 0.077, BF to NS: *p* < 0.001, *ph*^2^ = 0.763, MM to NS: *p* < 0.001, *ph*^2^ = 0.192, 2.5: BF to MM: *p* < 0.001, *ph*^2^ = 0.687, BF to NS: *p* < 0.001, *ph*^2^ = 0.874, MM to NS: *p* < 0.001, *ph*^2^ = 0.156) (Table 1). For the stride length, the results showed significant increases from BF to MM to NS in both speeds (2.0: BF to MM: *p* < 0.001, *ph*^2^ = 0.063, BF to NS: *p* < 0.001, *ph*^2^ = 0.190, MM to NS: *p* < 0.001, *ph*^2^ = 0.047, 2.5: BF to MM: *p* < 0.001, *ph*^2^ = 0.159, BF to NS: *p* < 0.001, *ph*^2^ = 0.284, MM to NS: *p* < 0.001, *ph*^2^ = 0.025).

**Table 1.** Stride length, footstrike angle and JCF for the hip, knee and ankle joint as mean ±standard deviation for all subjects (n = 16) during shoe conditions (barefoot = BF, minimalist shoe = MM, normal shoe = NS) and two velocities. Significance is indicated using asterisks, one = *p* < 0.05, two = *p* < 0.01, three = *p* < 0.001. + = significant difference BF to MM, * = significant difference BF to NS, ◊= significant difference MM to NS

|  |  | BF | MM | NS | p |
| --- | --- | --- | --- | --- | --- |
| stride length [m] | 2.0 m/s | 1.45 ± 0.09 | 1.49 ± 0.09 | 1.52 ± 0.09 | +++ , *** , ◇◇◇ |
|  | 2.5 m/s | 1.77 ± 0.11 | 1.84 ± 0.10 | 1.86 ± 0.10 | +++ , *** , ◇◇◇ |
| footstrike angle [°] | 2.0 m/s | 5.28 ± 2.53 | 7.91 ± 4.18 | 8.68 ± 3.88 | +++ , *** , ◇◇◇ |
|  | 2.5 m/s | 6.13 ± 2.82 | 8.18 ± 2.87 | 8.75 ± 2.47 | +++ , *** , ◇◇◇ |
| hip JCF [BW] | 2.0 m/s | 9.11 ± 1.88 | 8.98 ± 1.52 | 8.56 ± 1.43 | +++ , *** , ◇◇◇ |
|  | 2.5 m/s | 9.67 ± 1.59 | 9.64 ± 1.88 | 9.15 ± 1.84 | +++ , *** , ◇◇◇ |
| knee JCF [BW] | 2.0 m/s | 9.42 ± 1.47 | 9.45 ± 1.47 | 9.40 ± 1.30 |  |
|  | 2.5 m/s | 10.28 ± 1.68 | 10.48 ± 1.50 | 10.47 ± 1.72 | ++ , * |
| ankle JCF [BW] | 2.0 m/s | 5.58 ± 0.50 | 5.70 ± 0.38 | 6.01 ± 0.54 | +++ , *** , ◇◇◇ |
|  | 2.5 m/s | 6.03 ± 0.53 | 6.18 ± 0.48 | 6.59 ± 0.76 | +++ , *** , ◇◇◇ |
| hip flexion [°] | 2.0 m/s | -12.42 ± 5.93 | -13.88 ± 6.79 | -14.40 ± 5.53 | +++ , *** , ◇◇◇ |
|  | 2.5 m/s | -12.84 ± 5.77 | -13.92 ± 6.06 | -13.41 ± 5.88 | +++ , *** , ◇◇◇ |
| knee flexion [°] | 2.0 m/s | 38.32 ± 3.99 | 40.12 ± 3.30 | 41.22 ± 3.49 | +++ , *** , ◇◇◇ |
|  | 2.5 m/s | 39.18 ± 3.26 | 40.67 ± 3.60 | 41.49 ± 3.56 | +++ , *** , ◇◇◇ |
| ankle flexion [°] | 2.0 m/s | -23.56 ± 3.08 | -23.70 ± 2.92 | -23.51 ± 2.69 | +++ , ◇ |
|  | 2.5 m/s | -23.11 ± 3.00 | -23.26 ± 2.95 | -22.63 ± 2.57 | +++ , *** , ◇◇◇ |

### Kinematics

For the hip and knee, an increased flexion was observed in shod running. For the ankle, the observed changes were inconsistent. The hip flexion showed a significant increase at 2 m*/*s (2.0: BF to MM: *p* < 0.001, *ph*^2^ = 0.009, BF to NS: *p* < 0.001, *ph*^2^ = 0.015, MM to NS: *p* < 0.001, *ph*^2^ = 0.056) (Figure 2). For 2.5 m/s the hip flexion also increased between BF and both shoe conditions (MM and NS), but decreased significantly between MM and NS (2.5: BF to MM: *p* < 0.001, *ph*^2^ = 0.017, BF to NS: *p* < 0.001, *ph*^2^ = 0.186, MM to NS: *p* < 0.001, *ph*^2^ = 0.001).

The knee flexion showed a dependence on footwear over both speeds, with an increase in knee flexion from BF to MM to NS per speed (2.0: BF to MM: *p* < 0.001, *ph*^2^ = 0.504, BF to NS: *p* < 0.001, *ph*^2^ = 0.761, MM to NS: *p* < 0.001, *ph*^2^ = 0.251, 2.5: BF to MM: *p* < 0.001, *ph*^2^ = 0.610, BF to NS: *p* < 0.001, *ph*^2^ = 0.878, MM to NS: *p* < 0.001, *ph*^2^ = 0.221).

For the ankle, in 2.0 m/s the dorsalflexion increased between BF and MM (*p* < 0.001, *ph*^2^ < 0.001) and decreased between MM and NS (*p* = 0.016, *ph*^2^ < 0.001). Between BF and NS no significant change was found. In 2.5 m/s the dorsalflexion increased between BF and MM (*p* < 0.001, *ph*^2^ = 0.003) and decreased between BF and NS and MM and NS (BF to NS: *p* < 0.001, *ph*^2^ = 0.002, MM to NS: *p* < 0.001, *ph*^2^ = 0.007).

### Kinetics

The results showed an increase in JCFs for the ankle and a decrease for the hip when comparing barefoot running and shod running. For the knee, the JCF showed a slight increase at the higher speed.

The hip JCFs decreased significantly in both speeds from from BF to MM (2.0: *p* < 0.001, *ph*^2^ = 0.001, 2.5: *p* = 0.001, *ph*^2^ < 0.001), from BF to NS (2.0: *p* < 0.001, *ph*^2^ = 0.018, 2.5: *p* < 0.001, *ph*^2^ = 0.026), and from MM to NS (2.0: *p* < 0.001, *ph*^2^ = 0.010, 2.5: *p* < 0.001, *ph*^2^ = 0.013) (Table 1).

The effect of shoes on the knee JCF was only significant for the higher speed (2.5: BF to MM: *p* = 0.001, *ph*^2^ < 0.001), BF to NS: *p* = 0.010, *ph*^2^ < 0.001)). For MM to NS at the higher speed as well as for all comparisons at the lower speed, no significant changes were found.

The ankle JCFs in both speeds increased from BF to MM (2.0: *p* < 0.001, *ph*^2^ = 0.005, 2.5: *p* < 0.001, *ph*^2^ = 0.005), from BF to NS (2.0: *p* < 0.001, *ph*^2^ = 0.059, 2.5: *p* < 0.001, *ph*^2^ = 0.118) and MM to NS (2.0: *p* < 0.001, *ph*^2^ = 0.031, 2.5: *p* < 0.001, *ph*^2^ = 0.057). Figure 1 shows all peak JCFs for the hip, knee and ankle.

**Figure 1.**
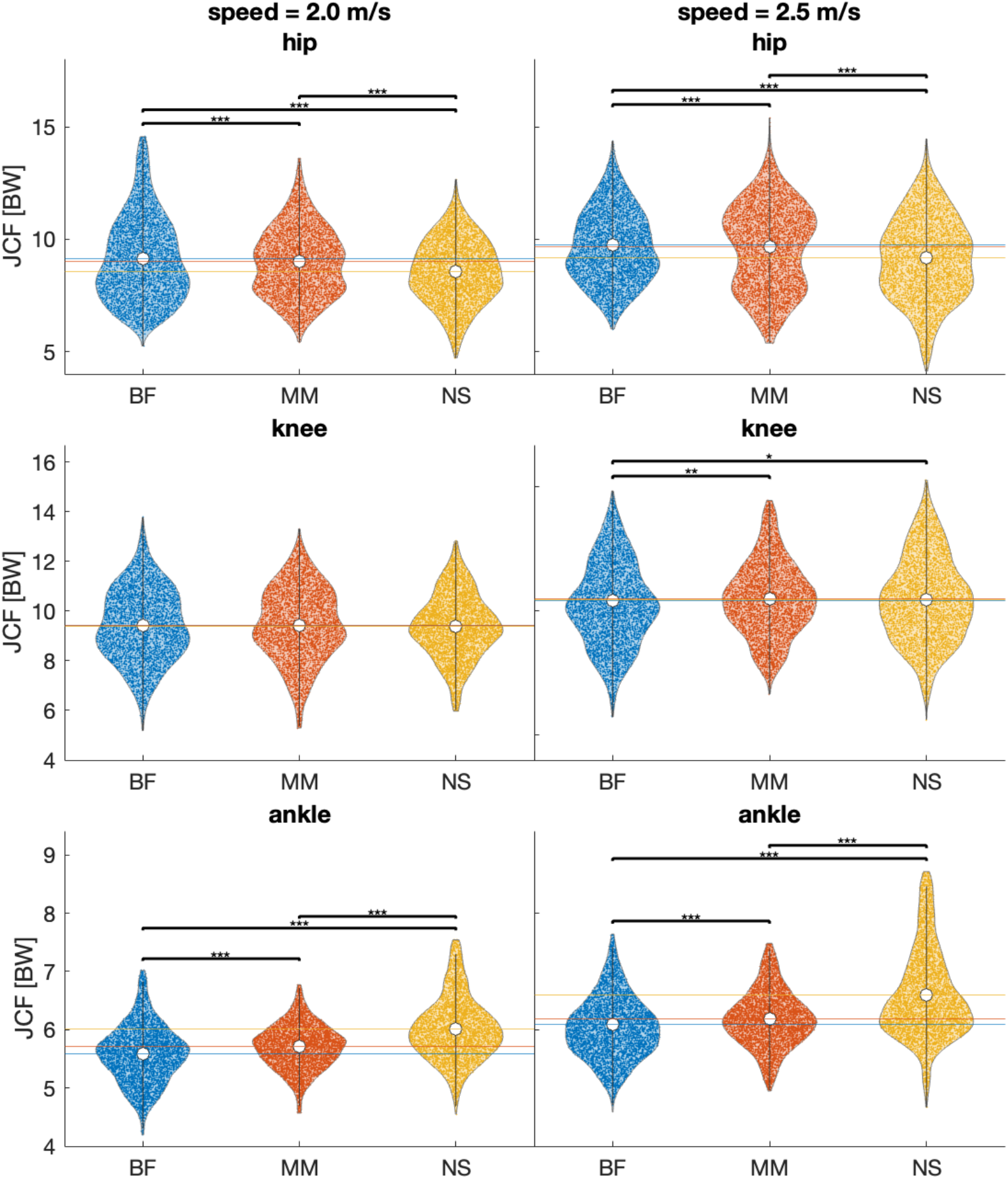
Peak joint contact force (JCF) of all steps of all participants per footwear condition (BF = barefoot, MM = minimal shoes, NS = normal shoes) in both speeds (2.0 m/s left, 2.5 m/s right) and for all joints (hip: top, knee: middle, ankle: bottom). Each data point represents one stride. Additionally, the median values are indicated by a white dot, and the interquartile range as a grey bar. Significance between conditions is indicated using asterisks: ^*^ *p* < 0.05, ^**^ *p* < 0.01, ^***^ *p* < 0.001

**Figure 2.**
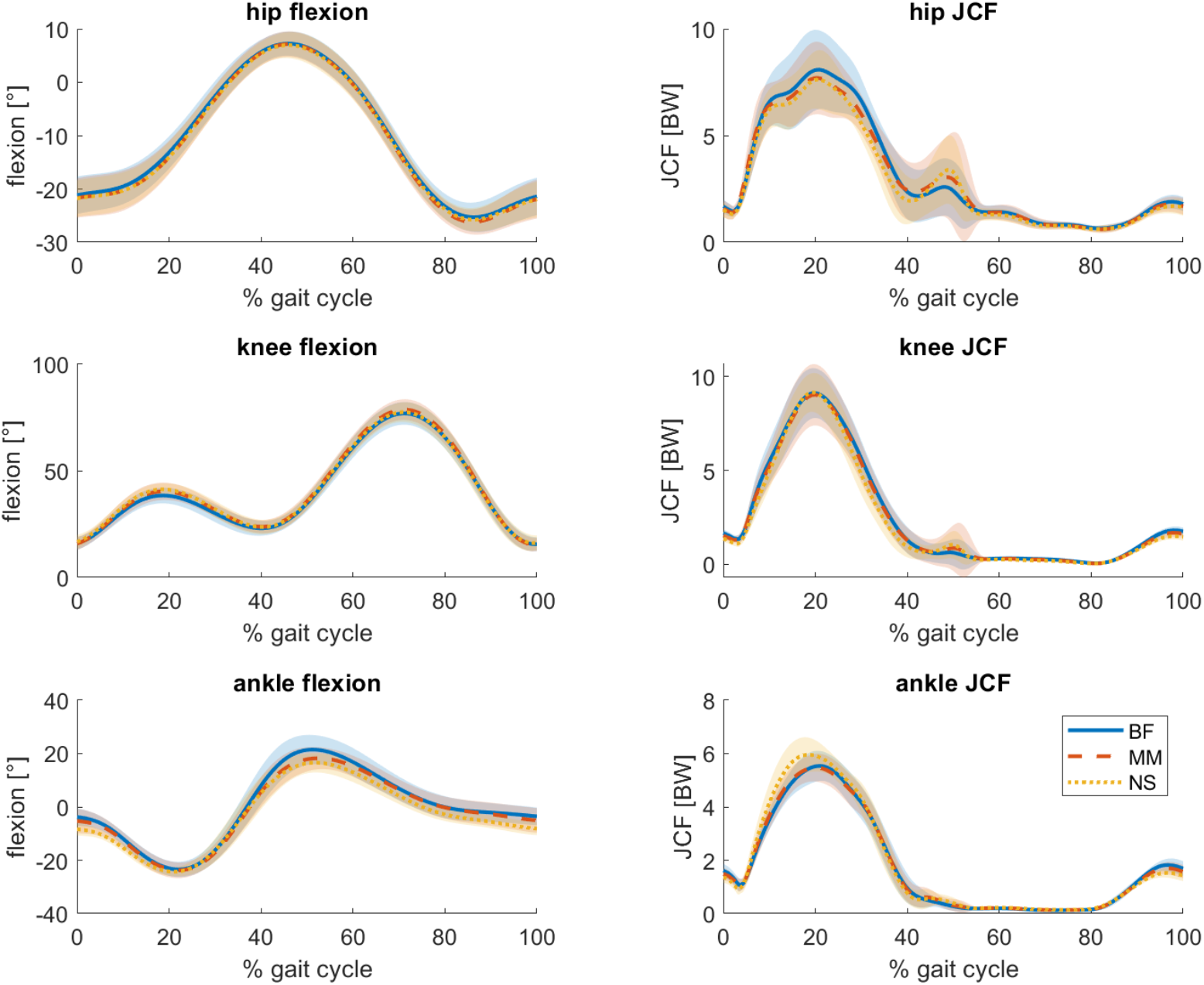
Joint flexion angles (left) and joint contact forces (right) per footwear condition as a function of the stride cycle during walking at 2.0 m/s. Values are averaged over all steps of all subjects (n=16) for the hip (top), knee (middle) and ankle(bottom) per condition. Barefoot running (BF) is indicated with a solid blue line, minimal shoes (MM) with a dashed orange, and normal shoes (NS) with a dotted yellow line. An angle of zero indicates the joint angles in the anatomical position. Positive angles indicate increased extension at the hip, increased flexion at the knee and increased plantarflexion at the ankle. Shaded areas indicate the standard deviation around the mean

### Individual responses

Individual changes between the different types of footwear conditions varied greatly (See Figure 3). While median responses over all participants were often close to zero, the changes per subject were often much larger. For the hip, the highest range can be found for the change from BF and MM in 2.0 m*/*s, where the individual changes range from −18.55% up to 27.93%. At the knee, the highest range is between the BF and the NS conditions at 2.0 m/s, where changes from −21.76% up until 23.13% were observed. At the ankle, the highest range was also observed from BF to NS at 2.0 m/s with −18.79 % the largest decrease and 33.93% the largest increase. As indicated in table 2, most of those changes are statistically significant regardless if they increased or decreased, with sometimes even large effect sizes.

**Table 2.**
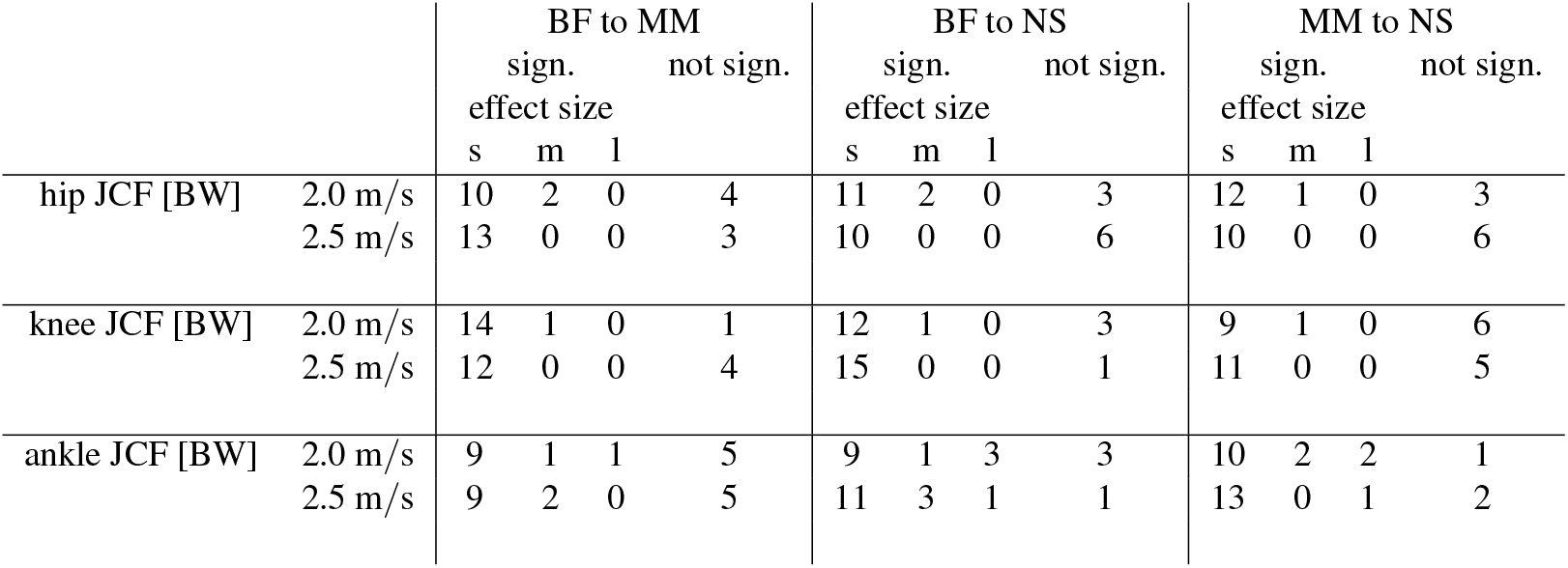
The number of subjects who showed statistically significant (sign.) or non-significant changes in joint contact forces (JCF) from one footwear condition to the other (BF: barefoot, MM: minimal, NS: normal shoes) in the two different speeds. Additionally, the effect size for the significant change is given as small (s), medium (m) or large (l) according to Cohen’s rule of thumb, which resulted in small effects being smaller than 0.035, large effects being higher than 0.1 and medium being between those two.^50^.

**Figure 3.**
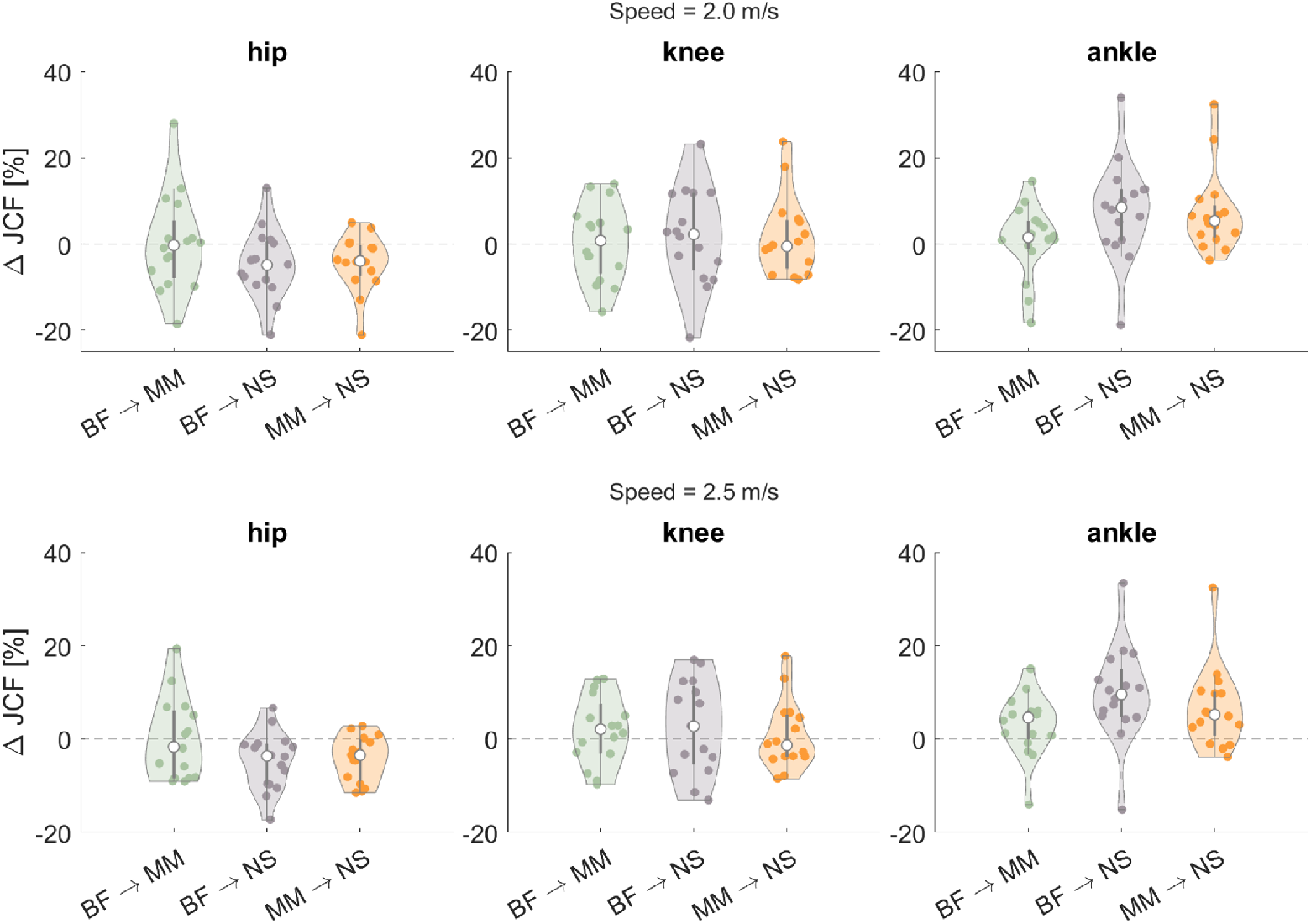
Relative change in the average peak JCFs between conditions (BF: barefoot, MM: minimal, NS: normal shoes) for the hip (left), knee (middle) and ankle (right) for running at 2.0 m/s (top) and 2.5 m/s (bottom). Change is calculated relative to the first-mentioned condition, e.g., (MM-BF)/BF. The median response over all participants is indicated with a white dot and interquartile ranges with a grey bar.

## Discussion

The aim of this study was to compare the joint contact forces of the hip, knee and ankle between different footwear conditions at two running velocities. Our hypothesis that peak JCFs are higher during shod running than during barefoot running could partially be confirmed: we found significantly increased JCFs in shod versus barefoot running at both speeds for the ankle.

At the knee, however, we only found a significant increase at 2.5 m*/*s, but not at 2.0 m*/*s. At the hip, we found significantly decreased JCFs, which went against our hypothesis. Alongside the JCFs, the kinematics of the three joints and stride parameters were analysed. We found significant changes in kinematics in all three joints. In the hip and knee, the flexion increased from barefoot running to shod running. In the ankle, the dorsalflexion increased from barefoot to minimalist and decreased from there to the normal shoe. Additionally, we observed increased stride lengths and footstrike angles in both shod conditions compared to barefoot running.

Our finding that the footstrike angle and stride length increase from the barefoot to the shod conditions is in line with current literature^6–9^and might be an adaption mechanism to the loss of shoe-induced damping in barefoot running. During running in conditions with less cushioning, such as barefoot running, the impact of the ground undergoes less shoe-induced damping than when running with more cushioned shoes. Switching to a less steep footstrike angle is a well-known adaptation mechanism to deal with this higher impact^2,6,8,12,22,27^, as it produces a body internal dampening by the foot arch and calf^10^. The smaller stride lengths that we observed in barefoot running fit in this picture, as smaller stride lengths have been shown to be connected to less steep footstrike angles^24^. Therefore, reducing the footstrike angle and stride length is a natural way of increasing the damping of the ground reaction forces in the body, when the shoe-induced damping is decreased.

These larger stride lengths provide a potential reason for the larger ankle JCF observed in the shod conditions versus barefoot running. With bigger stride lengths, the leg is placed further from the centre of mass^2^, resulting in a larger moment arm for the ground reaction forces^23,28^. This necessitates larger muscle forces to be generated^2,28^, leading to increased JCFs^15,15,20,24,33–36^. This idea is supported by findings from Bowersock et al., who found that larger step lengths are accompanied by larger JCFs^51^. While this theory agrees with our findings for the ankle JCFs it does not explain that we found an opposite effect at the hip and no consistent effect at the knee. Therefore, the larger stride length might explain some of the differences found in the JCF, but it seems like this effect is influenced by other factors as well.

As the amount of cushioning in the MM lies somewhere in between that of the BF and NS conditions, we expected the same to be the case for the recorded values of all investigated variables, with either a consecutive increase or decrease from BF to MM to NS. However, while mostly true, this was not always the case. In some cases, for example in the knee JCF, we see an increase from BF to MM, but no change from MM to NS. This effect can also be seen for some of the joint angles (Table 1). In our hypothesis, we suspected that the lack of cushioning of the shoes would result in a change of joint angles, as it has been shown that cushioning affects the joint angles^14,15^. However, even though the cushioning is a lot different between the MM and the NS, we do not see those matching changes in flexion angles or JCFs. This indicates that the differences in shoe effects are not solely down to the amount of cushioning.

While most of the group changes are statistically significant due to the large number of steps, the effect sizes tend to be small for the majority of significant variables. Only the ankle occasionally shows a medium or large effect size, e.g. between BF and NS at 2.5m/s. As mentioned above, this joint is most effected by stride parameters, which could in turn lead to stronger effects sizes. In total, however, the small effect sizes indicate that the found group differences might not be practically relevant, and we, therefore, want to emphasise that care should be taken when interpreting the group results.

Closer investigation indicates that the small group differences may, in part, be due to high inter-individual variability (Figure 3). Similar to earlier findings from literature^40–42^, we found large differences in the way individual subjects responded to the changes in footwear with some showing large effects to changes in footwear, while others show no significant reponse. We do not only find large differences in the magnitude of the change but, more importantly, also in the direction of change. For each joint, between 2 and 7 subjects show a change in JCF that is opposite to the main effect. The range of responses is quite large, e.g., responses in ankle JCFs ranged from −18% to +33% for a change from barefoot to conventional shod running at 2.5 m/s. This also shows in the effect sizes which range from very small up to large effects for individual subjects. This indicates that the effects of footwear interventions are highly individualistic and might be relevant for an individual person. Therefore, generalised statements about the influence of footwear should be made with care.

In order to gain some more insight into individual response patterns, we performed a correlation analysis of the responses to footwear changes between all variables. By comparing the percentage change of the JCF at the hip, knee and ankle each with the percentage change in the other outcome variables, we wanted to see if subjects who react in a specific manner to a footwear condition responded similarly in the other variables. This analysis yielded no significant correlation (see Supplementary Figure S.3). This means, that every subject has an individual reaction between different conditions for every investigated variable. We also checked if the results for the correlation analysis changed if the changes of the JCF were correlated with the absolute JCF values. Here as well, we found no significant correlations between the variables. Based on our current methods, we could find no indications for distinct groups of responders. However, future studies might be able to find more conclusive results by using suitable clustering methods.

When interpreting our results, it should be taken into account that only habitual rearfoot strikers were included for participation. Different strike patterns are accompanied by differences in the kinematics and kinetics^2,8,22,24,52,53^. Additionally, subjects were instructed to stay in a rearfoot strike pattern in all conditions. This could have changed their strike pattern, especially in barefoot running, where it has been shown that people adopt a more midfoot to forefoot strike pattern^2,6,8,22,27,54^. However, there are also studies that show that, while subjects run with a more plantarflexed footstrike angle, this change is limited, as they do not switch to a complete forefoot strike pattern^12,27^. In our study subjects did run with a more plantarflexed footstrike in barefoot, but did not switch to a forefoot strike. Therefore, we are able to distinguish between the effects of footwear and strike pattern changes and can conclude that the changes that we found are indeed because of different footwear conditions and not because of a different strike pattern.

The reported JCFs have been calculated using a musculoskeletal model, rather than recorded *in vivo*. As the Myonardo is an inverse dynamic model, it is not an exact representation of the human body and it comes with known model limitations^55,56^. However, this kind of method is needed in order to asses joint forces without the use of invasive measures^55–57^. Additionally, the Myonardo has been validated for the knee during squatting and walking movements on data recorded using instrumented prostheses data^44^. We performed an additional validation, which can be found in the Appendix, where calculated hip JCFs are compared to JCFs recorded with an instrumented hip endoprosthesis during a single leg stance task^45^(see Supplementary Figure S.1). This is a movement which is fairly comparable to the (one-legged) stance phase during running.

Our study shows that, when switching from barefoot to shod running, hip JCFs decrease and ankle JCFs increase, while findings on the knee JCFs were inconsistent. While significant, the relative changes in the forces were quite small. Therefore, it is uncertain how clinically relevant they are. The main reason for the small effects are the large inter-individual differences. Subjects did not only differ in the magnitude of the response, but also showed opposite responses to the same footwear interventions. The individual differences were much larger than the overall response, and thus, more likely to be clinically relevant. Our findings hereby indicate that footwear interventions should be made on a case-by-case basis. Future research should focus on identifying defining runners’ characteristics for the different types of responder.

## Data availability

Data is available upon request to the corresponding author Lena Kloock.

## Acknowledgements

We thank Predimo GmbH for providing the Myonardo software.

## Author contributions statement

L.K. conceived and conducted the experiments, analysed the results, and wrote the manuscript A.A. conceived and conducted the experiments, analysed the results, and wrote the manuscript M.d.G conceived the experiments, analysed the results and wrote the manuscript M.G. conceived and conducted the experiments K.B. analysed the results and wrote the manuscript H.W. conceived the experiments and analysed the results All authors reviewed the manuscript.

