## Supplementary material for "Joint contact forces during barefoot, minimal and conventional shod running are highly individual"

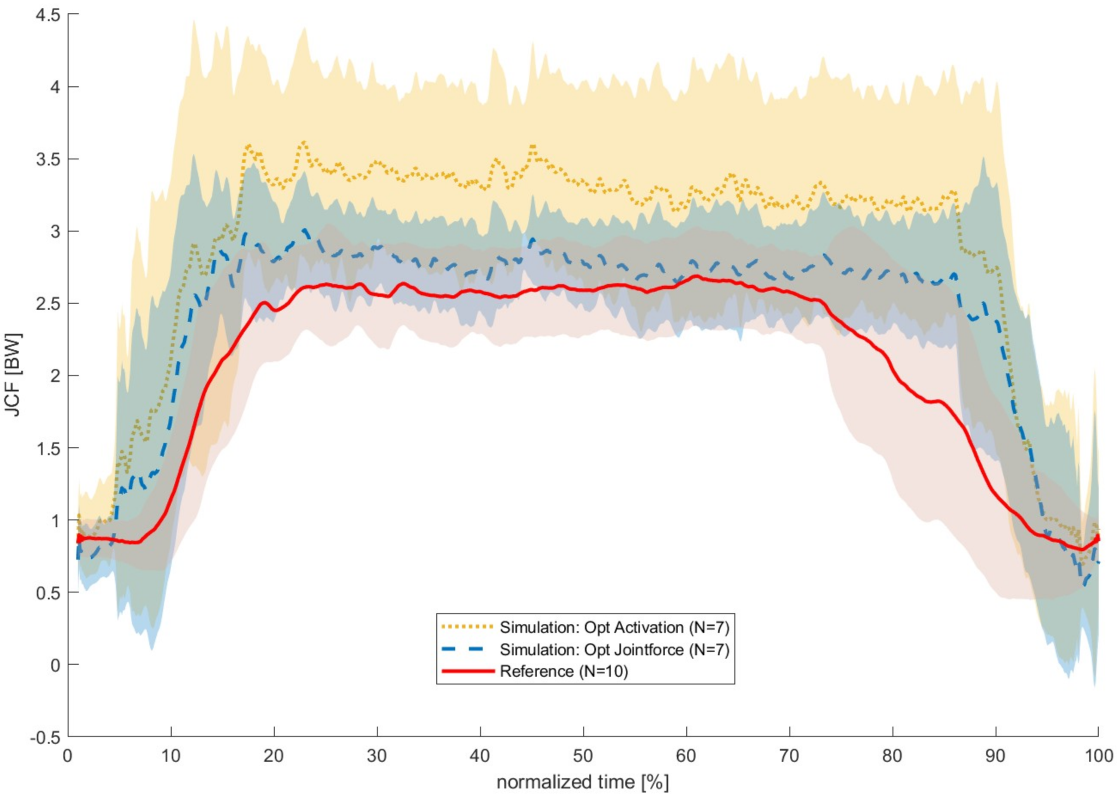


**Figure S.1.** Comparison of hip JCFs measured with an instrumented endoprosthesis (red) to rearranged simulated absolute joint contact forces (JCFs) of the hip with the Myonardo® (blue) during a one-legged stance as mean ± standard deviation.The movement data that was used for the calculation of the JCFs was collected with the same protocol as the reference data[1], but with younger, able bodies subjects without a previous hip replacement. In order to ensure a better comparison and to account for the impaired subjects of the validated data, the rearranged data was simulated using a muscle optimization to minimise the joint force, which was deemed appropriate for subjects with an impaired hip (opt: joint force). Additionally the rearranged data was simulated in the same manner that was used in the study, which is more appropriate for the subject group: a minimisation of the muscle activation[2]. All values were normalised to bodyweight and the time was normalised to fit all participants. Please note, that the reference data is from subjects aged 52 up until 68, while the subjects for the rearranged data were between 23 and 35, and it would thus be expected for them to have slightly higher hip contact forces[3]. The reference data was extracted from the database OrthoLoad[1] for 10 subjects (files, appended with 'one_leg_stance': H1L_060511_1_024, H2R_150811_1_024, H3L_141111_2_075, H4L_270_112_2_036, H5L_050412_1_037, H6R_201112_1_036, H7R_191112_1_038, H8L_240413_1_036, H9L_301013_1_029, H10R_300114_1_032).

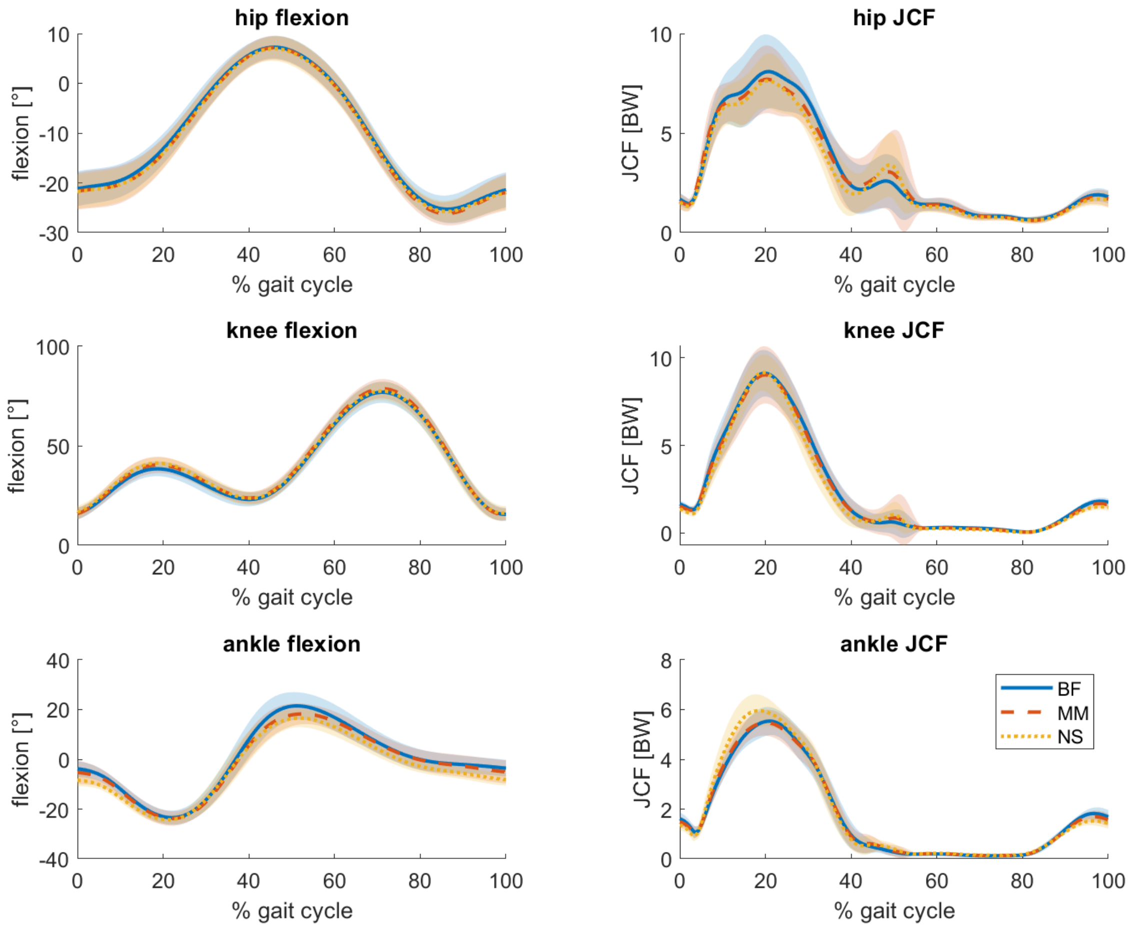


**Figure S.2.** Joint flexion angles (left) and joint contact forces (right) per footwear condition as a function of the stride cycle during running at 2.0 m/s. Values are averaged over all steps of all subjects (n=16) for the hip (top), knee (middle) and ankle (bottom) per condition. Barefoot running (BF) is indicated with a solid blue line, minimal shoes (MM) with a dashed orange, and normal shoes (NS) with a dotted yellow line. An angle of zero indicates the joint angles in the anatomical position. Positive angles indicate increased extension at the hip, increased flexion at the knee and increased plantarflexion at the ankle. Shaded areas indicate the standard deviation around the mean.

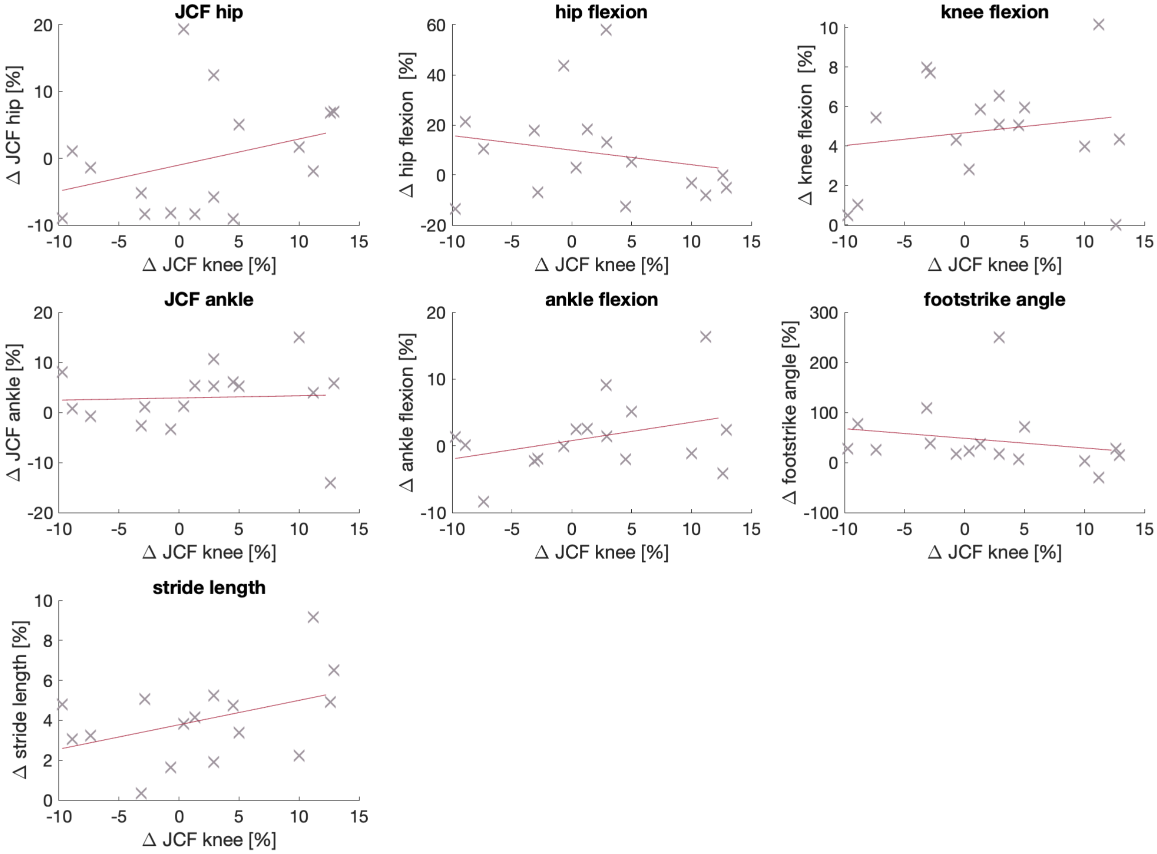


**Figure S.3.** Linear regression between the percent change in joint contact force (JCFs) at the knee and the percent change in all other variables from BF to MM at 2.5 m/s. Each cross represents the response of one individual. Correlation analysis was performed for these variables and yielded no significant correlations

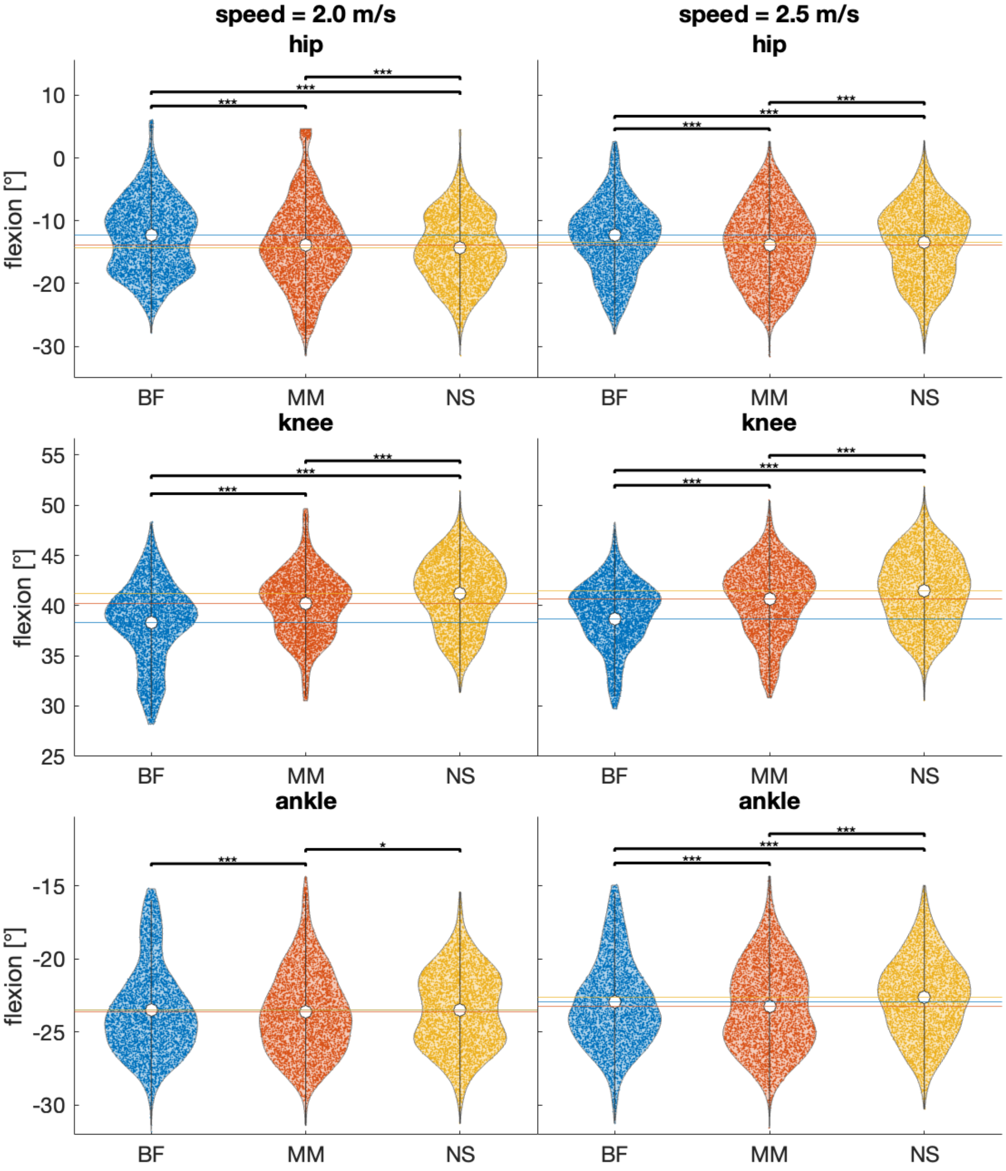


**Figure S.4.** Flexion angles at peak joint contact force (JCF) of all steps of all participants per footwear condition (BF = barefoot, MM = minimal shoes, NS = normal shoes) in both speeds (2.0 m/s left, 2.5 m/s right) and for all joints (hip: top, knee: middle, ankle: bottom). Each data point represents one stride. Additionally, the median values are indicated by a white dot, and the interquartile range as a grey bar. Significance between conditions is indicated using asterisks: $^{*}p < 0.05$, $^{**}p < 0.01$, $^{***}p < 0.001$

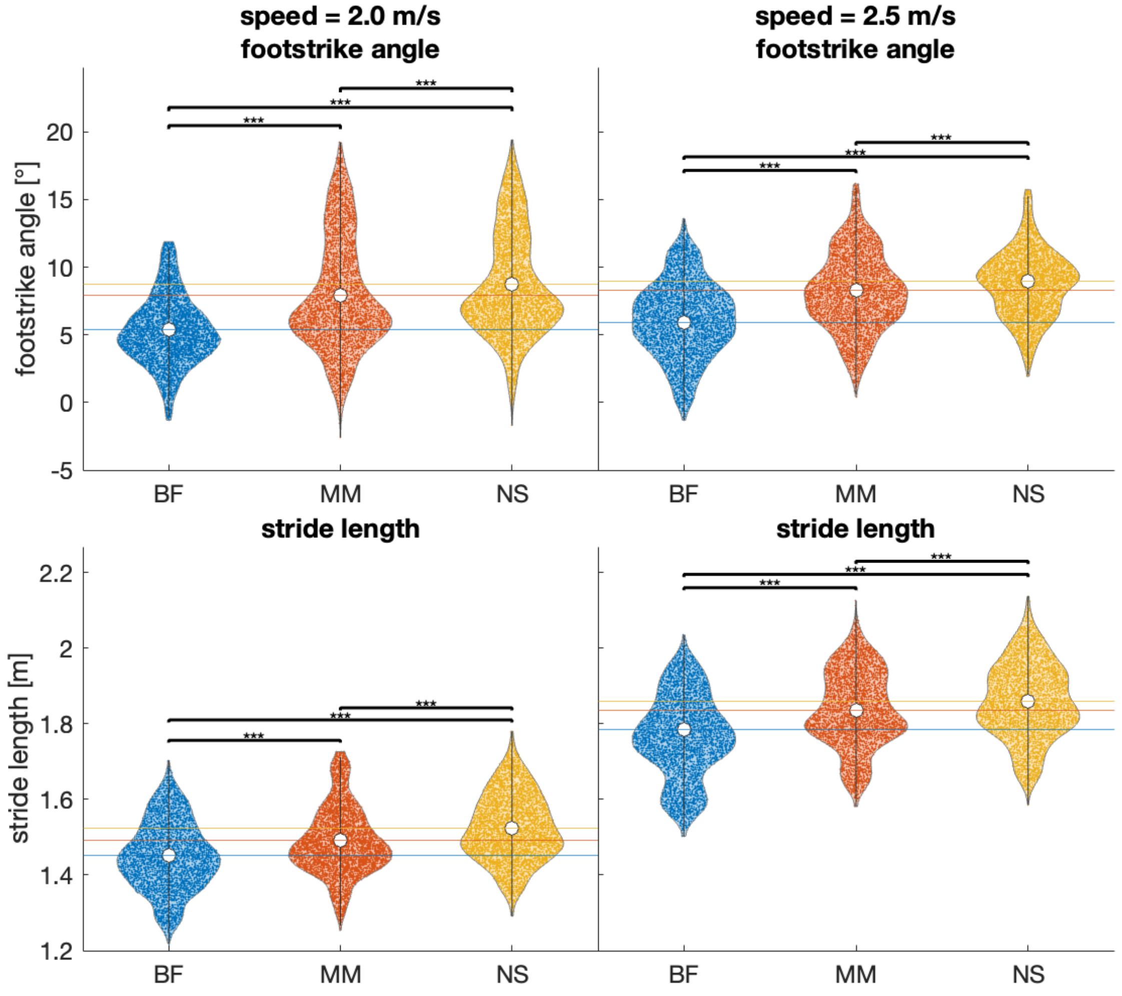


**Figure S.5.** Footstrike angle (top) and stride length (bottom) of all steps of all participants per footwear condition (BF = barefoot, MM = minimal shoes, NS = normal shoes) in both speeds (2.0 m/s left, 2.5 m/s right). Each data point represents one stride. Additionally, the median values are indicated by a white dot, and the interquartile range as a grey bar. Significance between conditions is indicated using asterisks: $^{*}p < 0.05$, $^{**}p < 0.01$, $^{***}p < 0.001$
